# The impact of Covid-19 vaccination on the Italian healthcare system: a scenario analysis

**DOI:** 10.1101/2021.06.08.21258561

**Authors:** Marcellusi Andrea, Fabiano Gianluca, Sciattella Paolo, Andreoni Massimo, Francesco S Mennini

## Abstract

**Introduction:** The objective of this study is to estimate the effects of the national immunisation strategy for Covid-19 in Italy on the national healthcare system.

**Methods:** An epidemiological scenario analysis was developed in order to simulate the impact of the Covid-19 pandemic on the Italian national healthcare system in 2021. Hospitalisations, ICU admissions and death rates were modelled based on 2020 data. We forecast the impact of the introduction of a primary prevention strategy on the national healthcare system by considering vaccine efficacy, availability of doses and potential population coverage over time.

**Results:** In the absence of immunisation, between 57,000 and 63,000 additional deaths are forecast in 2021. Based on the assumptions underlying the two epidemiological scenarios from the 2020 data, our model predicts that cumulative hospital admissions in 2021 will range from 3.4 to 3.9 million. The deployment of vaccine immunisation has the potential to control the evolution of 2021 infections and avoid from 60 to 67 percent of deaths compared to not vaccinating.

**Conclusions:** In order to inform Italian policymakers on delivering a mass vaccination programme, this study highlights and detects some key factors that must be controlled to ensure that immunisation targets will be met in reasonable time.

## 1. Introduction

The race to develop Covid-19 vaccines has occurred at an unprecedented pace. Nine months after the World Health Organization (WHO) announced the Covid-19 outbreak a global pandemic, the UK became the first nation to adminster the vaccine developed by Pfizer-BioNtech to people most at risk from the SARS CoV-2 coronavirus. Two other vaccine candidates, one developed by the US-based company Moderna and another by the English Oxford-AstraZeneca partnership, had been rolled out by the first quarter of of 2021. Several other promising vaccine trials are also underway.

The total production capacity of the three vaccines is estimated at 5.3 billion doses for 2021, which could cover between 2.6 and 3.1 billion people, approximately one-third of the world population. The European Union, together with five other high-income countries, have pre-ordered approximately half of these doses, although their citizens account for only around 13% of the global population [1].

Yet despite concerted efforts to secure most of the doses initially available, not everyone in developed nations will get the vaccine straight away. European countries have granted access to Covid-19 vaccines based on population size, and governments are following prioritisaton strategies to cover those most at risk first. In Italy, the immunisation plan gives priority to healthworkers, and subsequently to population groups most at risk, e.g. those older than 80 years of age and with comorbidities [2].

The effective deployment of vaccination strategies is subject to many factors that can potentially hasten or delay the achievement of herd immunity. These include the number of doses secured by each country, the speed at which the vaccines will be distributed by manufacturers, the speed at which these will be administered by health authorities, and the efficacy in reducing the risks of developing severe symptoms. Moreover, despite the fact that vaccines have been proved to be safe and effective against the development of clinical disease, there is no published data on whether and to what extent they could prevent virus transmission or on the duration of the induced protection, potentially affecting the logistics of immunisation, for example, if boosters will be needed [3]. Finally, building public trust through clear communication of the benefits of vaccination is fundamental in achieving high levels of vaccination take-up by the general population [4].

As Covid-19 infections are still spreading across most EU nations, health authorities are called to deal with the spread of infection while deploying initial vaccination strategies. In Italy, the incidence of Covid-19 infections among older patients is one of the factors which is putting the Italian National Health System (SSN) under constant pressure. Despite efforts made to increase hospital infrastructure, Italy faces a comparatively lower intensive care unit (ICU) capacity than those of other European countries, and is characterised by regional healthcare systems that differ widely in terms of hospital organisation, equipment and medical workforce [5]. Therefore, introducing Covid-19 vaccination has the potential to avoid hospital capacity saturation and progressively allow the release of regional restrictions such as lockdowns and limitations on economic activity. Optimising the outcomes of immunisation is a key factor in meeting disease reduction goals and limiting the impact of Covid-19 on healthcare systems and mortality.

The objective of this study is to forecast the effects of the Italian national immunisation strategy in terms of mortality rates and demand on the healthcare service. An epidemiological scenario analysis was developed in order to simulate the impact of the Covid-19 pandemic on the Italian national healthcare system in 2021. We forecast the impact of the introduction of a primary prevention strategy on the national healthcare system by considering vaccine efficacy, availability of doses, and potential population coverage over time. Different scenarions were developed in terms of avoided hospital admissions and deaths for 2021.

## 2. Methods

### 2.1 Epidemiological scenarios

Based on the available data from the Covid-19 pandemic curve in 2020, a number of assumptions were made in order to forecast the progression of the pandemic in 2021 and the expected effects of immunisation.

We identified three key moments in the 2020 records of infections (See Appendix Figure 1), as follows:

– A first key moment, the ‘first wave’, was defined by the pattern of infections recorded between February and April 2020. During this time, a peak of around 29,000 hospitalisations was recorded, with an average of 950 daily deaths (Table 1). The response by the Italian government was to introduce a series of progressive restrictions in order to delay the diffusion of coronavirus, including limitations in individual mobility and the closure of social, cultural, economic and industrial activities [6, 7]. These restrictions resulted in a decrease of Covid-19 cases and deaths until the end of March (see Appendix Figure 1).

**Table 1.**
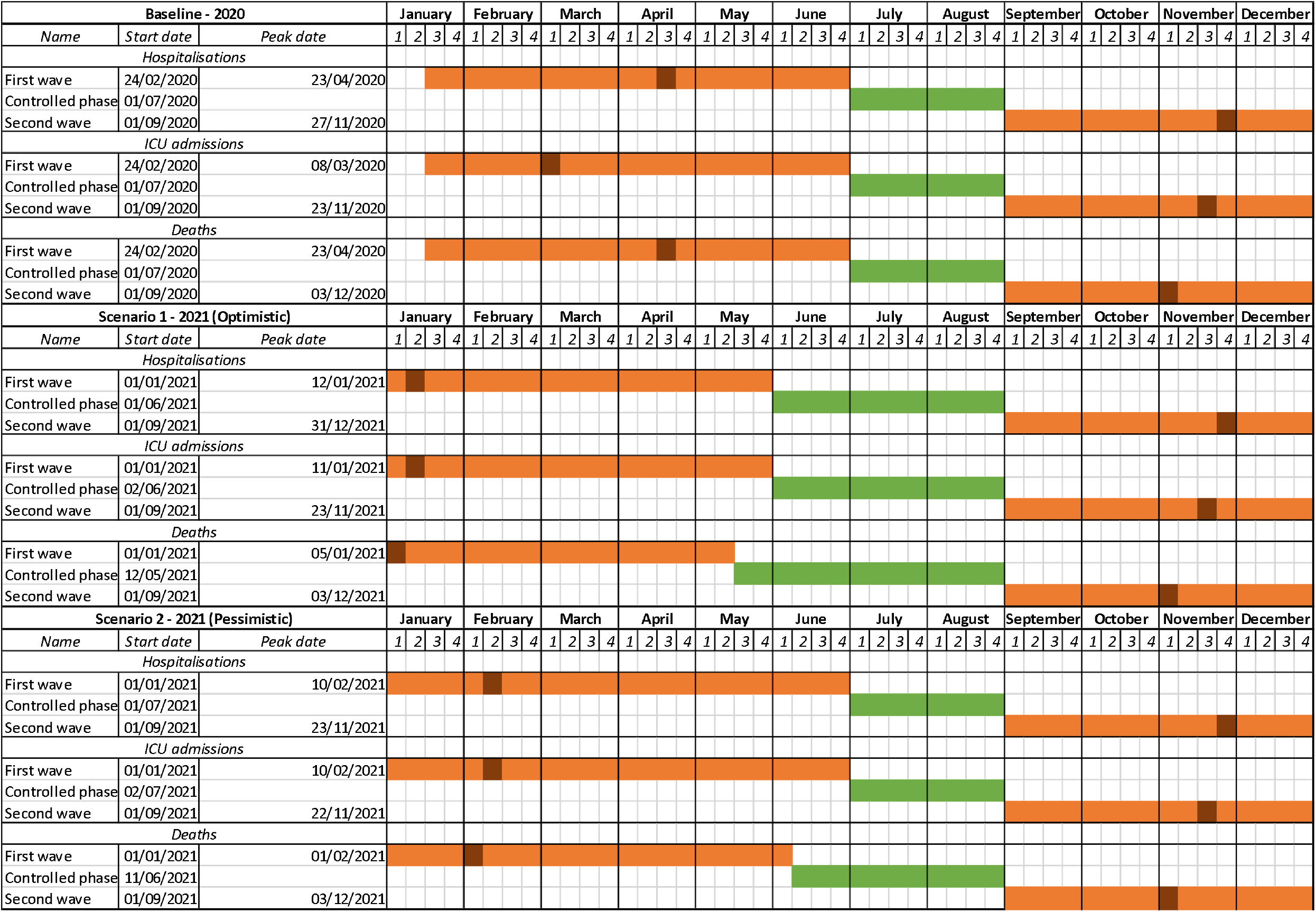
Epidemiological scenarios and time assumptions.
– The second key moment, the ‘controlled phase’, was the period between May and August 2020. In this time frame, a minimum of 713 hospitalisations and 3 daily deaths due to Covid-19 were recorded during the second half of July. During this period the health authorithies removed some restrictions to allow for free mobility between regions, and for some economic and social activities. The number of daily infections remained constant over time until September 2020.
– A third key moment, the ‘second wave’, pertained to the period between September and December 2020. During this time, Italy was hit by a second wave of infections, which was recorded as more intense and deadly than the first. Over 34,500 hospitalisations and 990 daily deaths were registerd in Italy at the end of November (see Appendix Figure 1). In contrast to that of the second wave of infections, the Italian government adopted a new containment strategy [8] valid between 6^th^ November and 3^rd^ December 2020, which divided Italy into three tiers (yellow, orange and red) of increased restrictions. This approach was updated on a weekly basis, and regions were allocated to one of the three tiers based on their most recent epidemiologic data (e.g. the number of infected people and free hospital beds in the previous week).

In order to predict the expected curve for 2021 without vaccination, our model replicates key moments recorded in 2020 by operating a shift in time based on the latest available infection rates at the time of the writing (February 2021). The most recent available data was matched with 2020 infection records and the corresponding points in time were identified. Specifically we identified timeframes when, in 2020, the pandemic curve showed similar values as those reported at the time of writing. Consequently, two scenarios were built:

– The first scenario (optimistic), was built on the assumption that the number of recorded cases was similar to those registered during the first wave, when the trend was decreasing.
– The second scenario (pessimistic), was built on the expectation of a growing trend of infection, similarly to what recorded in late 2020.

The assumptions employed to build Scenarios 1 and 2 are reported in Table 1, which shows the start and peak dates predicted for each key moment in 2021 and the corresponding shifted dates from 2020.

### 2.2 Data sources

Data was retrieved from the Italian National Institute of Statistics (ISTAT) and the Italian National Institute of Health (ISS) websites (https://www.epicentro.iss.it/coronavirus/sars-cov-2-sorveglianza-dati). Epidemiological data on the SARS-CoV-2 epidemic in Italy in terms of hospital admissions, access to ICU and deaths covering the period from 21^st^ February 2020 to 31^st^ January 2021 was downloaded. Age-stratified deaths rates were used to calculate the distribution of Covid-19-related hospitalisations and ICU admissions. Covid-19 events were classified by age group: <60, 60-79, and ≥80 years old (Supplementary Table 1). Risk of events were calculated as the ratio between the number of hospitalisations, ICU admissions and deaths in each age class and the number of resident population within the same age group.

### 2.3 Model parameters

A set of parameters was used to inform the estimation model and forecast the effect of immunisation on the 2021 pandemic curve.

#### 2.3.1. Vaccination scenarios

The first parameter was defined by the number of subjects to be vaccinated (with two doses) out of the total eligible population in each quarter. Given the initial reduced availability of vaccines in the first quarter of 2021, a prioritised approach was planned by Italian health authorities in which health workers and a specific at-risk population (those over 80 years old) were due to be vaccinated first [9]. Subsequently, vaccines were set to be administered to those with comorbidities and those aged between 60 and 79. Once these groups are protected, the national strategy is to deploy vaccines to the general population [9] (See Appendix Table 1).

Three vaccination approaches are employed in the model, based on the 2021 target dates for vaccinating 75% of the general population, as set by the Italian immunisation plan:

– A *base-case* approach was built on the expert opinion elicited from medical doctors at the Policlinico of University of Tor Vergata in Rome, which is one of 321 vaccination centres designated by the Italian health ministry as part of the national immunisation plan. In this approach, the model assumes that the target of immunising 75% of the population will be achieved between the third and fourth quarters of 2021. Specifically, the base-case approach assumes that 8% of the population would be vaccinated by March 2021, 30% by June 2021, 70% by September 2021, and 90% of the population before the end of 2021 (See Appendix Table 2).
– An *optimised* approach was built on the information reported in the national immunisation plan. In this scenario, the maximum number of available doses, as reported by the document, was employed. Therefore, this model assumes that all those doses that are planned to be available in each quarter will be administered to the population (Appendix). Under these assumptions, the target immunisation coverage (75%) would be achieved between the second and third quarters of 2021.
– Lastly, an approach with *minimum* vaccination coverage was built by considering the minimum level of vaccination coverage that is reported in the National Covid-19 vaccination program (Appendix). In this case, the target coverage would be achieved at the end of the fourth quarter.

#### 2.3.2 Stocks

The number of doses that are planned to be delivered in each quarter were derived from the national immunisation plan. In our model we assumed that all the doses would be delivered by the end of 2021 [9]. However, by operating on the coverage parameter, we also accounted for delays that could occur in the delivery of doses. At the time of writing, a delay of 60% of doses for the AstraZeneca vaccine was considered for the first quarter [10].

#### 2.3.3 Effectiveness

An explorative literature review was conducted in order to identify randomised clinical trial (RCT) studies that were conducted by vaccine producers. Effectiveness was defined in terms of the number of reduced symptoms between the vaccinated arm (experimental group) versus the placebo arm (control group) (Appendix). RCT effectiveness values were then employed to calculate risk ratios by age class (<60, 60-79 and ≥80 years old).

The estimated reduction in Covid-related events have been extrapolated considering the vaccinated and unvaccinated population in each scenario. The reduction rates of Covid-related events 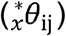 were applied to the daily event risk for hospitalisations, ICU admissions and deaths by applying the following formula:

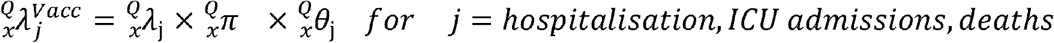

In the formula, 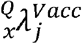 represents the daily event risk during the period Q by age class *x* in the simulation where Covid-19 vaccination is considered, 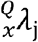 represents the daily event risk assumed for each quarter Q by age class without vaccination, 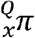 is the percentage of patients that will be vaccinated and 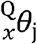 is the reduction rate of Covid-related events estimated by the model [11].

## 3. Results

In the absence of immunisation, between 57,000 and 63,000 additional deaths are forecast in 2021 (Table 2). Based on the assumptions underlying the two epidemiological scenarios from the 2020 data, the model predicts that cumulative hospital admissions will range from 3.4 to 3.9 million in 2021. Consequently, the pressure on intensive care units is projected to comprise between 351,000 and 414,000 admissions throughout the year (Table 2). The overall estimated impact of the Covid-19 pandemic in 2021, in the absence of vaccination, is reported in Figure 1.

**Table 2.** Number of estimated cases by scenario and coverage simulation – Italy 1^st^ January 2021-31^st^ December 2021.

| Epidemiological scenario | Vaccination approach | Cases |  |  | % of Avoided cases |  |  |
| --- | --- | --- | --- | --- | --- | --- | --- |
|  |  | Hospitalisations | ICU admissions | Cumulative deaths | Hospitalisations | ICU admissions | Cumulative deaths |
| Optimistic Scenario 1 | **No Vaccination** | 3,433,663 | 351,914 | 57,748 |  |  |  |
|  | Base-case | 1,499,230 | 154,000 | 19,974 | 56.3% | 56.2% | 65.4% |
|  | Optimised | 1,345,552 | 136,846 | 18,975 | 60.8% | 61.1% | 67.1% |
|  | Minimum | 1,614,274 | 162,395 | 21,292 | 53.0% | 53.9% | 63.1% |
| Pessimistic Scenario 2 | **No Vaccination** | 3,951,503 | 414,670 | 63,200 |  |  |  |
|  | Base-case | 1,908,316 | 209,578 | 23,168 | 51.7% | 49.5% | 63.3% |
|  | Optimised | 1,693,989 | 182,040 | 21,703 | 57.1% | 56.1% | 65.7% |
|  | Minimum | 2,058,444 | 221,161 | 25,090 | 47.9% | 46.7% | 60.3% |

**Figure 1.**
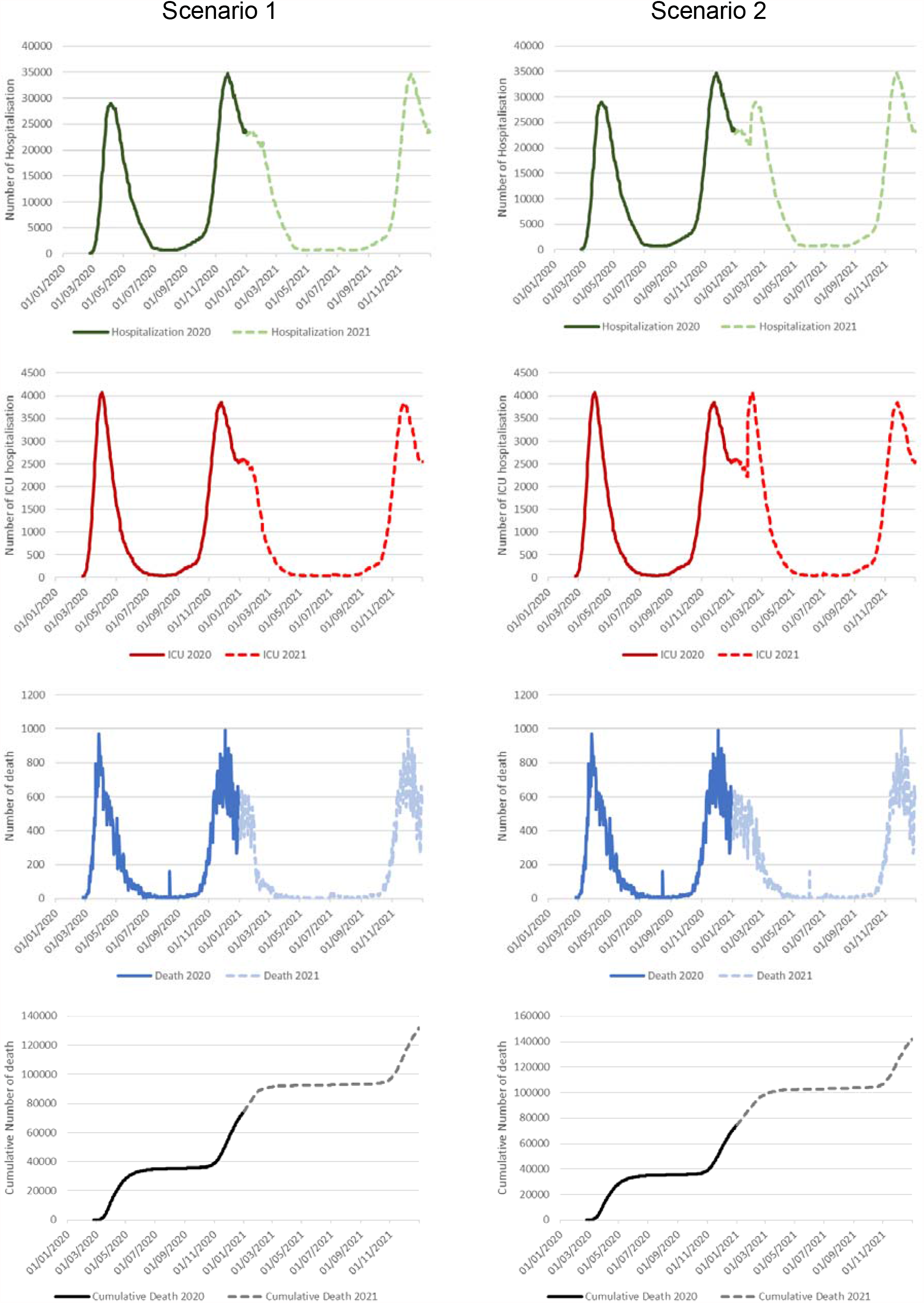
Covid-related events without vaccination by scenario – Italy 1^st^ January 2020-31^st^ December 2021.

In Table 2, the number of cases that could potentially be avoided with vaccination is reported based on the assumptions of the pessimistic and optimistic epidemiological scenarios, and the simulations of the different coverage rates that could be achieved throughrough 2021.

The deployment of vaccine immunisation has the potential to control the evolution of 2021 infections and avoid hospital over-utilisation and excess deaths. Overall, the model predicts a reduction of 60-67 percent deaths compared to not vaccinating (Table 2). Furthermore, the pressure on the Italian healthcare system is estimated to decrease 48-60 percent in terms of reduced demand on hospital and ICU services.

In the case of a optimistic epidemiological outlook (Scenario 1) and an optimal deployment of the vaccination campaign, cumulative deaths are estimated at 19,000 in 2021. In this scenario, there woud be a decrease of 65.4% in the number of deaths compared to the no-vaccination simulation. Furthermore, the projected 1.3 million hospitalisations (compared to 3.4 million that are expected without the vaccination programme) represents a much reduced pressure on the healthcare system. ICU admissions of 136,000 are predicted throughout 2021.

In the case of a pessimistic epidemiological scenario and an inefficient deployment of the immunisation plan, our model forecasts that 25,000 deaths caused by coronavirus in 2021. Hospital and ICU admissions are expected to show 2.5 million and 221 thousands respectively, which corresponds to a reduction of 47.9 and 46.7 percent from the no-vaccination scenario.

All the epidemiological and vaccination scenarios are showed in Figure 2 in terms of expected deaths, hospitalisations and ICU admissions over time in 2021.

**Figure 2.**
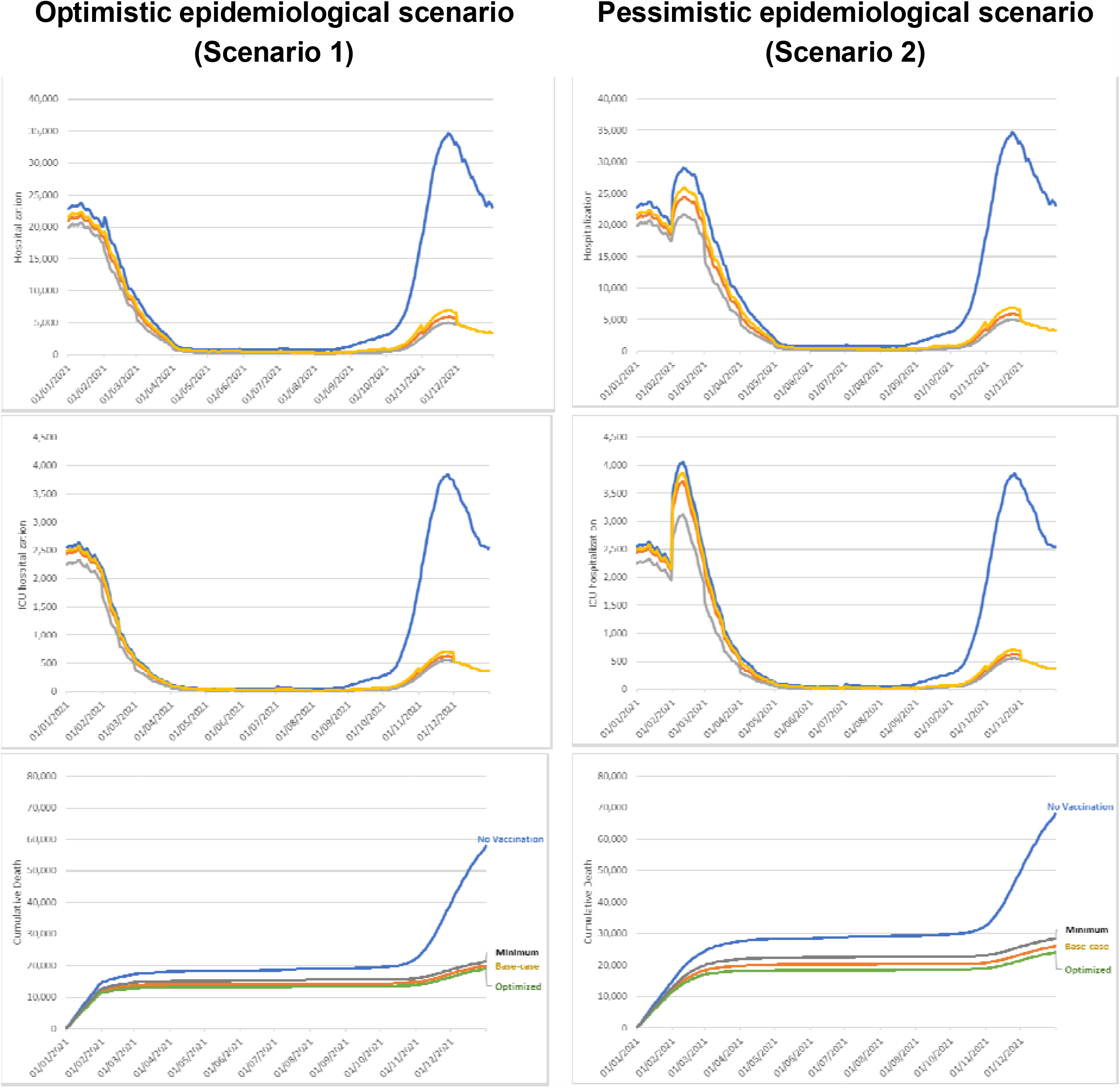
Estimated deaths, hospitalisations and access to ICU by epidemiological and vaccination scenarios in 2021.

## 4. Discussion

Actors in the biopharmaceutical arena have made extraordinary efforts to develop effective vaccine solutions in record time. Yet the race to make these vaccines available to populations has only just begun, and there are several challenges for health authorities at local levels.

With the aim of informing Italian policymakers’ decision-making and actions to bring vaccinination to the general population, this study has highlighted and detected some key factors that must be controlled to ensure that immunisation targets will be achieved in areasonable time.

Our model highlights that the availability of doses in each quarter of 2021, the efficacy levels and the efficiency of health facilities in the administration are key factors that would allow for greater control over the spread of infections and the number of deaths. In particular, securing enough doses to protect the most fragile population groups and the professionals most at risk is the main driver for driving down infection and death rates in the short term.

An increasing number of authors have tried to forecast the evolution of the Covid-19 pandemic in Italy [12-14]. However, most of these forecasts have tried to estimate the overall number of new Covid-19 cases. To our knowledge, this is the first analysis that works on a hypothetical estimation of hospitalisations, ICU admissions and Covid-related deaths. Despite the simplicity of the forecast, no complex statistical model was used and our model tries to inform decision-makers in a timely and appropriate manner. The model’s ability to examine uncertainty parameters (e.g. pandemic wave evolution and peak) allows for the evaluation of a large number of plausible scenarios. Moreover, the analysis represents a first attempt to evaluate the primary prevention effects of the Covid-19 pandemic in Italy. As with other vaccination strategies [15], this model considers the main aspects of a vaccination approach (including vaccine efficacy, availability of doses and potential population coverage over time), and simulates minimum and maximum assumptions for assessing uncertainty.

Covid-19 is an unequal killer [16] with a significant impact on disability and life expectancy [17]. Central to the Italian crisis and the medical response were the healthcare workers who were exposed to Covid-19 without having any perception of what they were facing during the first pandemic wave [18, 19]. Because of their vitally important role in the crisis, at the time of writing (February 2021) the Italian healthcare system have vaccinated over 1.2 million people in Italy, over 90% of whom are health workers [20]. The number of individuals vaccinated during the first month of the immunisation programme is in line with the our model’s estimation and confirms the efforts made by the Italian government. As of January 2021, the Italian healthcare system has administered over 91% of the doses that were distributed by the vaccine producers [20]. This demonstrates that the Italian healthcare system can perform efficiently in the administration of vaccines. Although, the ability to maintain an efficient vaccine distribution and administration system across the whole of 2021 represents the first challenge for vaccination success and for obtaining the results estimated in the optimised scenario of our model. Of course, the availability of additional vaccine doses also plays a significant part in reaching vaccination coverage targets faster.

This study has some limitations that also open opportunities for future research. First, the model did not include any epidemiological simulations and herd immunity was not considered in the analysis. Consequently, the indirect protection caused by the increasing size of the vaccinated population is a factor that has the potential to increase the effects of immunisation more quickly than is forecast in this model. Second, several assumptions were made based on the data and the knowledge accumulated in 2020. The data informing our model is being constantly updated and therefore, some of the assumptions used may require an update. However, the model will be constantly calibrated and updated with new data and therefore it represents a potential tool that policymakers may find useful in the control of the pandemic. Lastly, the efficacy shown by vaccines needs to be further demonstrated in the real world, in particular regarding the duration of the protection and whether vaccinated people can be infectious.

## 5. Conclusion

Covid-19 has put the Italian economic and health systems under immense pressure. Restrictions have been implemented to control the spread of the virus and to make sure that the health system was able to bear the additional demands caused by the pandemic. While controlling the spread of infections and protecting the national health system from collapse, restrictions were introduced at an incredibly high cost to the economy.

The discovery of effective vaccines now has the potential to bring new tools to control the spread of the virus, and contribute towards a solution for the economy versus health trade-off. Vaccines are key tools to control the pressure on healthcare system and, at the same time, allow the return to commercial and social activities.

This study aimed to analyse the primary effects of introducing Covid-19 vaccination on the Italian national healthcare system. We developed an estimation model to forecast the potential benefits of immunisation in terms of reduced mortality, hospitalisations and ICU admissions. By doing so, this work provides an initial estimate of the reliefs that vaccinating fragile and at-risk populations in anticipation of the whole population could bring to the Italian health and economic systems.

The results of our study also highlight that the benefits from vaccination are subject to specific conditions. In particular, we show the contrasting impact of a less efficient and slower vaccination campaign, especially in the context of a still severe epidemiological scenario in which the virus still spreads quickly across the entire population. Furthermore, we illustrate that although increasing the number of available doses could boost vaccination intake, this is also dependent on infrastructure capacity and the allocation of sufficient workforce, which must be planned and managed accordingly. Moreover, the results of this work can be used by policymakers to build public trust, which is itself a fundamental condition to reach satisfactory coverage rates in the general population. In the light of these conditions, this study is a first attempt to quantify health benefits from vaccinations with the potential to inform and drive further economic analysis. The model has the potential to be constantly calibrated and updated with new information, and therefore to act as a policy tool for supporting decision-making.

## Data Availability

All the data are available in the text of the main document

## Author Approval

all authors have seen and approved the manuscript.

