## Supplementary Material for "The impact of Covid-19 vaccination on the Italian healthcare system: a scenario analysis"

Figure 1 - Age distribution of Covid-19-related evenst – Italy 1st January 2020-16th January 2021

| 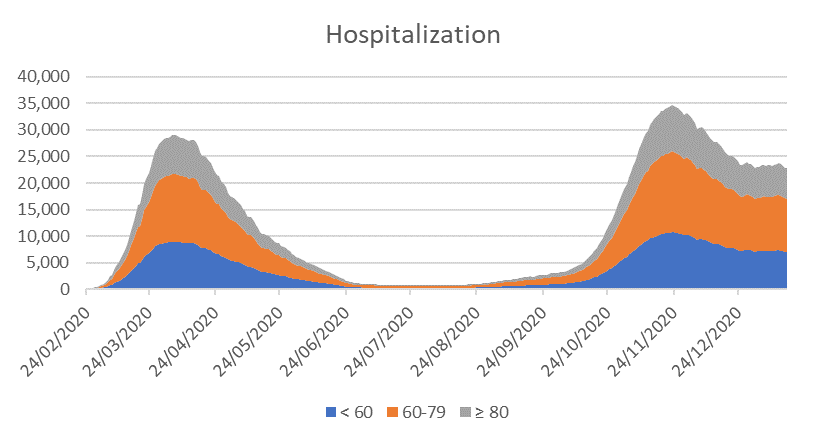 | 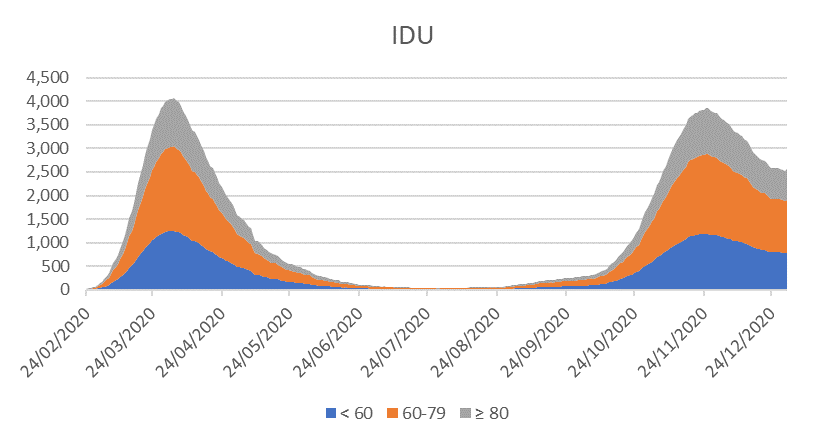 |
| --- | --- |
| 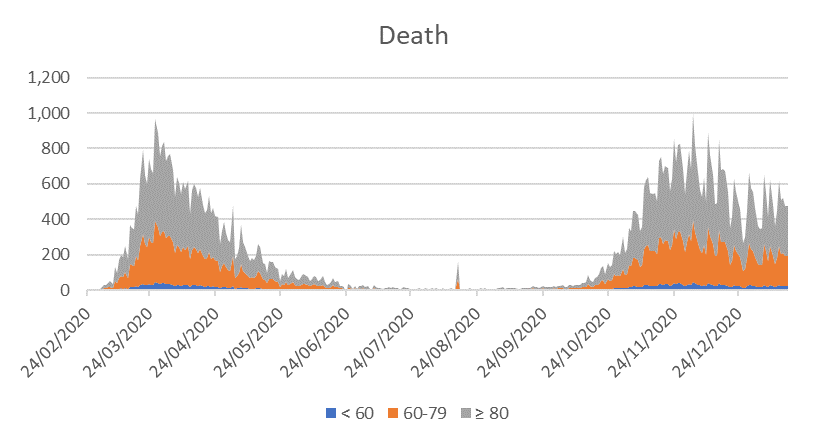 | 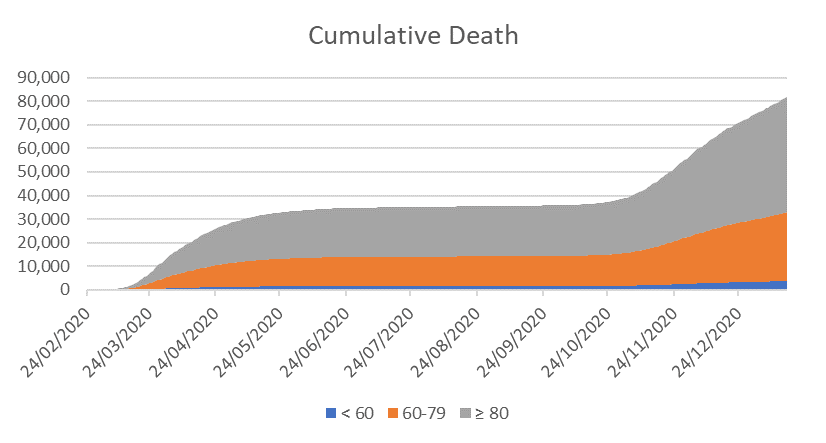 |

Data from Italian National Institute of Statistics (ISTAT) and the Italian National Institute of Health (ISS) websites (<https://www.epicentro.iss.it/coronavirus/sars-cov-2-sorveglianza-dati>).

Age distribution for hospitalisations and IDU admissions were estimated considering a sample of 996 Covid-19 hospitalisations recorded in Policlinico Tor Vergata Hospital between 2^nd^ March 2020 and 27^th^ December 2020 (Table 1).

Table 1 - Distribution of Covid-19-related events by age class in Italy

| **Age Class** | **Hospitalisations^[1]^** | | **ICU Admissions** | | **Deaths^[2]^** | |
| --- | --- | --- | --- | --- | --- | --- |
|  | **N** | **%** | **N** | **%** | **N** | **%** |
| < 60 | 308 | 31% | 49 | 22% | 1,907 | 11% |
| 60-79 | 437 | 44% | 152 | 67% | 14,834 | 51% |
| 80 + | 251 | 25% | 26 | 11% | 24,989 | 38% |
| **Total** | **996** | **100%** | **227** | **100%** | **41,730** | **100%** |

Table 2 - Approaches to vaccination coverage, by age category and time.

| a) **Base-case** | | | | | |
| --- | --- | --- | --- | --- | --- |
| *Age category* | Population | Q1 | Q2 | Q3 | Q4 |
| ≥80 | 4,442,048 | 90.0% | 0.0% | 0.0% | 0.0% |
| 60-79 | 13,432,005 | 5.1% | 84.9% | 0.0% | 0.0% |
| <60 | 42,370,586 | 0.0% | 4.1% | 58.1% | 27.8% |
| **Overall coverage rate** | **60,244,639** | **7.8%** | **29.6%** | **70.5%** | **90.0%** |
| b) **Optimised** | | | | | |
| *Age category* | Population | Q1 | Q2 | Q3 | Q4 |
| ≥80 | 4,442,048 | 90.0% | 0.0% | 0.0% | 0.0% |
| 60-79 | 13,432,005 | 39.4% | 50.6% | 0.0% | 0.0% |
| <60 | 42,370,586 | 0.0% | 51.5% | 38.5% | 0.0% |
| **Overall coverage rate** | **60,244,639** | **15.4%** | **62.9%** | **90.0%** | **90.0%** |
| c) **Minimum** | | | | | |
| *Age category* | Population | Q1 | Q2 | Q3 | Q4 |
| ≥80 | 4,442,048 | 67.8% | 22.2% | 0.0% | 0.0% |
| 60-79 | 13,432,005 | 0.0% | 37.5% | 52.5% | 0.0% |
| <60 | 42,370,586 | 0.0% | 0.0% | 33.1% | 56.9% |
| **Overall coverage rate** | **60,244,639** | **5.0%** | **15.0%** | **50.0%** | **90.0%** |

Table 3 - Estimated doses available (million) in Italy per quarter

|  | Q1 | Q2 | Q3 | Q4 | Total |
| --- | --- | --- | --- | --- | --- |
| Astrazeneca | 16,155,000* | 24,225,000 |  |  | 40,380,000 |
| Pfizer/BioNtech | 8,749,000 | 8,076,000 |  |  | 16,825,000 |
| J&J |  | 14,806,000 | 32,304,000 | 6,730,000 | 53,840,000 |
| Sanofi/GSK |  |  | 20,190,000 | 20,190,000 | 40,380,000 |
| Curevac | 2,019,000 | 5,384,000 | 6,730,000 | 8,076,000 | 22,209,000 |
| Moderna | 1,346,000 | 4,711,000 | 4,711,000 |  | 10,768,000 |
| Total | 28,269,000 | 57,202,000 | 63,935,000 | 34,996,000 | 184,402,000 |

Source: Ministry of Health, Strategic Vaccination anti-SARS-CoV-2/COVID-19 Plan

* A 60% reduction was considered in the model simulation

Figure 2 - Summary of available evidence on vaccination efficacy (last update 20/01/2021)

| **Vaccine** | **Ages Eligible** | **Actual Study Start Date** | **Estimated Study Completion Date** | **Efficacy primary end-point** | **Statistical Measure** | **Observed efficacy  (95% C.I.)** | **Source** |
| --- | --- | --- | --- | --- | --- | --- | --- |
| ASTRA | 18 - 130 | August 28, 2020 | February 21, 2023 | Efficacy of ChAdOx1 nCoV-19 vaccine against virologically confirmed, symptomatic COVID-19, defined as a NAAT-positive swab combined with at least one qualifying symptom (fever ≥37·8°C, cough, shortness of breath, or anosmia or ageusia) occurred ≥ 15 days post second dose of study intervention. | 100 x (1 - RR_adj_) | 70.4 (54.8 - 80.6) | 1 |
|  |  |  |  |  |  | 62.1 (41.0 - 75.7) |  |
|  |  |  |  |  |  | 90.0 (67.4 - 97.0) |  |
| PF/BT | ≥ 16 | April 29, 2020 | January 27, 2023 | Efficacy of BNT162b2 against confirmed Covid-19 with onset at least 7 days after the second dose in participants who had been without serologic or virologic evidence of SARS-CoV-2 infection up to 7 days after the second dose. | 100 × (1 − IRR) | 95.0 (90.3 - 97.6) | 2 |
| Moderna | ≥ 18 | July 27, 2020 | October 27, 2022 | Efficacy of the mRNA-1273 vaccine in preventing a first occurrence of symptomatic Covid-19 with onset at least 14 days after the second injection in the per-protocol population, among participants who were seronegative at baseline. | 100 x (1 - HR_adj_) | 94.1 (89.3 - 96.8) | 3 |
| J&J | ≥ 18 | September 7, 2020 | March 10, 2023 | Efficacy of Ad26.COV2.S in the prevention of first occurrence of Molecularly Confirmed Moderate to Severe/Critical Coronavirus Disease (COVID-19) with Seronegative Status [14 days post-vaccination (Day 15) to end of study]. |  |  | 4 |
| Sinovac | ≥ 18 | September 2021 | October 2021 | Incidence of Covid-19 cases after two-doses immunisation schedule [Time Frame: two weeks after second dose up to one year after first dose]. |  |  | 5 |
| Beijing / Elea | 18 - 85 | September 16, 2020 | December 1, 2021 | Incidence of Covid-19 cases after two doses of vaccination [Time Frame: 14 days after the full course of vaccination]. |  |  | 6 |
| Sputnik-V | ≥ 18 | September 7, 2020 | May 1, 2021 | Percentage of trial subjects with coronavirus disease 2019 (Covid-19) developed within 6 months after the first dose. |  |  | 7 |
| CanSino | ≥ 18 | September 15, 2020 | January 30, 2022 | Incidence of Covid-19 cases [Time Frame: day 28 to 12 months post-vaccination]. |  |  | 8 |
| Wuhan Provincial Center | ≥ 18 | July 16, 2020 | July 15, 2021 | To evaluate the protective effect 14 days after 2 doses of immunisation of preventing severe cases of SARS-CoV-2 pneumonia and deaths accompanied by Covid-19. |  |  | 9 |
| Novavax | 18 - 84 | September 28, 2020 | January 14, 2022 | Number of participants testing serologically negative for severe acute respiratory syndrome coronavirus 2 (SARS-CoV-2) at baseline, with first occurrence of positive (+) polymerase chain reaction (PCR)-confirmed SARS-CoV-2 illness with symptomatic mild, moderate, or severe Covid-19 with onset from Day 28 through the length of the study. |  |  | 10 |
| Medicago | ≥ 18 | November 19, 2020 | April 30, 2022 | First occurrence, in a subject, of laboratory-confirmed (virologic method) symptomatic SARS-CoV-2 infection [Time Frame: 14 days]. |  |  | 11 |
| Bharat Biotech | 18 - 99 | November 19, 2020 |  | To evaluate the efficacy of BBV152B to prevent Covid-19 based on the case definition for the secondary efficacy symptomatic endpoint [Day 42 to Month 12]. |  |  | 12 |
| Anhui Zhifei | ≥ 18 | December 16, 2020 | April 30, 2022 | The endpoint of efficacy study [Time Frame: 14 days to one year after whole vaccination]. |  |  | 13 |
| Curevac | ≥ 18 |  |  | Occurrence of first episodes of virologically-confirmed (RT-PCR positive) cases of Covid-19 of any severity meeting the case definition for the primary efficacy analysis. |  |  | 14 |
| AnGes, Inc. | ≥ 18 | November 23, 2020 | March 31, 2022 | Rate of SARS-CoV-2 positive and incidence rate of Covid-19 after the first vaccination [Time Frame: Week 1 through Week 53]. |  |  | 15 |
| Chinese Academy of Medical Sciences | ≥ 18 | December 2020 | March 2022 | The incidence of Covid-19 cases after two doses of vaccination [Time Frame: From 14 days after the second dose to 1 year after the second dose]. |  |  | 16 |
| Clover | ≥ 18 | December 2020 | July 2022 | Number of participants with a first occurrence of Covid-19 of any severity starting 14 days after second dose of SCB-2019 [Time Frame: Day 36 up to Day 389 (1 year after second dose)]. |  |  | 17 |

1. https://www.thelancet.com/journals/lancet/article/PIIS0140-6736(20)32661-1/fulltext
2. https://www.nejm.org/doi/full/10.1056/NEJMoa2034577
3. https://www.nejm.org/doi/full/10.1056/NEJMoa2035389
4. https://clinicaltrials.gov/ct2/show/study/NCT04505722?term=NCT04505722&draw=2&rank=1
5. https://clinicaltrials.gov/ct2/show/NCT04456595?term=vaccine&cond=COVID-19&phase=2&draw=2
6. https://clinicaltrials.gov/ct2/show/NCT04560881?term=vaccine&cond=covid-19&draw=2
7. https://clinicaltrials.gov/ct2/show/NCT04530396
8. https://clinicaltrials.gov/ct2/show/NCT04526990
9. http://www.chictr.org.cn/showprojen.aspx?proj=56651
10. https://clinicaltrials.gov/ct2/show/NCT04583995?term=Novavax&draw=2
11. https://clinicaltrials.gov/ct2/show/NCT04636697?term=Medicago&draw=2
12. http://ctri.nic.in/Clinicaltrials/pmaindet2.php?trialid=48057&EncHid=&userName=sars-cov-2%20vaccine
13. https://clinicaltrials.gov/ct2/show/NCT04646590
14. https://www.curevac.com/wp-content/uploads/2020/12/20201214-CureVac-HERALD-Clinical-Trial-Protocol-of-Phase-2b_3_CVnCoV.pdf
15. https://clinicaltrials.gov/ct2/show/NCT04655625
16. https://www.clinicaltrials.gov/ct2/show/NCT04659239

https://clinicaltrials.gov/ct2/show/NCT04672395

1. Derived from Policlinico Tor Vergata Hospital
2. Derived from recorded death by age – Italian Ministry of Health monitoring Data updated 11/11/2020
